# Forecasting COVID-19 impact in India using pandemic waves Nonlinear Growth Models

**DOI:** 10.1101/2020.03.30.20047803

**Authors:** Pavan Kumar, Ram Kumar Singh, Chintan Nanda, Himangshu Kalita, Shashikanta Patairiya, Yagya Datt Sharma, Meenu Rani, Akshaya Srikanth Bhagavathula

## Abstract

The ongoing pandemic of the coronavirus disease 2019 (COVID-19) started in China and devastated a vast majority of countries. In India, COVID-19 cases are steadily increasing since January 30, 2020, and the government-imposed lockdown across the country to curtail community transmission. COVID-19 forecasts have played an important role in capturing the probability of infection and the basic reproduction rate. In this study, we predicted some trajectories of trajectories associated with COVID-19 in the coming days in India using an Auto-regression integrated moving average model (ARIMA) and Richard’s model. By the end of April 2020, the incidence of new cases is predicted to be 5200 (95% CI: 4650 to 6002) through the ARIMA model versus be 6378 (95% CI: 4904 to 7851) Richard model. We estimated that there would be a total of 197 (95% CI: 118 to 277) deaths and drop down in the recovery rates will reach around 501 (95% CI: 245 to 758) by the end of April 2020. These estimates can help to strengthen the implementation of strategies to increase the health system capacity and enactment of social distancing measures all over India.

## Introduction

The ongoing pandemic of the coronavirus disease 2019 (COVID-19) started in China and devastated a vast majority of countries. Due to rapid pandemic potential and the absence of antiviral drugs and vaccines, this contagious COVID-19 disease has recorded thousands of deaths across the world [1]. COVID-19 placed tremendous strain on the health system and left dilemma with large case numbers. In India, COVID-19 cases are steadily increasing since January 30, 2020, and the government-imposed lockdown across the country to curtail community transmission.

Mathematical models are widely used to forecast the spreading of the disease and capture the probability of cases from susceptible to infected, and then to a recovery state or death. Many SIR models have been published or proposed online [2-5]. However, these models assume randomly mixed between all individuals in the given population. Nonlinear models/functions are more advanced methods that provide solution iteratively [6]. The iterative methods such as nonlinear regression include the modified Gauss-Newton method, gradient or steepest-descent method, multivariate secant or false position, and the Marquardt method [6,7]. With regards to COVID-19, forecasts have played an important role in capturing the probability of infection and the basic reproduction rate. No studies have used a specific nonlinear model to forecast the COVID-19 dynamics in India. Therefore, we generated 30 days forecast the dynamics of cumulative confirmed death and recovery of COVID-19 cases in India.

## Methods

We here used data from Johns Hopkins Corona Virus Resource Center (https://coronavirus.jhu.edu/), which reports very comparative data with cumulative cases for 170+ countries worldwide, including state or province wise for some of the special database cases. Here we have collected each day case data at given stipulated tie from the data of having date January 30, 2020, to March 28, 2020. Some date wise cases for confirmation of COVID-19 reporting cases along with total cumulative results of recovered cases and death cases are analyzed using statistical analysis. We here used Auto-regression integrated moving average model (ARIMA) and Richard’s model in the R-language platform. The new projected data is used up to April 29, 2020, for the creation of trajectory having projected score for the entire three cases reported-case confirmed, recovered, and death.

### Forecasting of projected prediction

Here we standardize all the models in a file format to detect the daily case for India country of available data. Reported data that is collected from entire sources is data that is considering from January 30, 2020, to March 28, 2020, so the projection trajectory will analyze up to April 29, 2020. The best fit analysis for India, which we included for analysis of the cumulative reported case and its upcoming necessity in the future as well as recovery and death cases.

### Statistical Models

We here used some statistical phenomenological models to detect and analyze the disease based trajectory model for prediction purposes. We precisely used four models to analyze the aggregate data set for time series analysis. This includes ARIMA and Richard’s model [8]. Another type of COVID-19, like SARS disease (Severe Acute Respiratory Syndrome), is analyzed without breaking the current situation and predicting the future perspective [9].

### Comments

Forecasting is exceptionally vital even to get the slightest result for multi variables consideration over public health factors, especially pandemic crises like COVID-19. In this case, forecast from single models is not enough for reliable results and prediction. Therefore, here we are using two different models integrated for time series analyses. Hence all the two different models will be discussed ahead [10].

Time series models provide a different and unique approach to time series forecasting. Basically, for the time series forecast, two approaches are widely used, i.e., exponential smoothing and Time Series Models like ARIMA and Richard’s. While exponential smoothing models are based on a description of the trend and seasonality in the data, Time Series, like ARIMA models, aims to describe the autocorrelations in the data.

### Stationarity and Differencing

A stationary time series where data properties do not depend on the time at which the series is observed. Therefore, time-series data with trends or with seasonality are not stationary as it will affect the value of the data at different times. In our study here, we are using machine learning tools for predicting the spread of COVID-19 in the future, so having a stationary time series data is very important for further predictable modeling. As we can see, the trend is followed by the variables used in our data for the victims affected by COVID-19. Therefore, to test whether the data is stationary or not becomes a very vital aspect of our research.

Here our time series data is based on the spread of the victims of the COVID-19 across India; hence, finding the correlation within the variable of recovering, confirmed and death cases will be crucial for the formation of the time series format for further modeling [11]. For another two trends like death and recovery cases is time lagging situation. So, to detect this analysis, we require some other statistical formulation and models. These are described, followed by how they influence to getting the results.

### Auto-Regressive Integrated Moving Average Model (ARIMA)

Now, here we combine differencing with auto-regression (AR) and Moving Average models (MA). The full model can be written as:

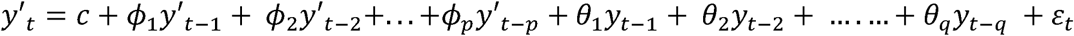

Where y′_t_ is the differenced series, the “predictors” on the right-hand side include both lagged values of y_t_ and lagged errors. We call this an ARIMA (p,d,q) model, where, p=order of the autoregressive part; d=degree of first differencing involved; q=order of the moving average part.

The Auto-Regressive Integrated Moving Average (ARIMA) is a model for times series data analysis based on their three-class components to manage stationary and non-stationary time datasets. The first component autoregressive (AR)(p) time series model for dependent observations for the forecast of future values. Even it is the function of dependent and dependent lag value and with any constant as white noise.

The moving average (MA)(q) model deals with white noise observations (past forecast errors) for the forecast for the future dependent value. Even it is the function of white noise and past white noise error. Both the combination will make the ARMA model, which deals with stationary data values. We are dealing with time-series non-stationary values, the data observed value means, and variance is not constant, so third component (integrating(I)(d)) was used to convert the observations using differencing series [12,13]. The differencing order two observation was used for the model forecast for COVID-19 case cumulative incidence, mortality, and recovery to avoid any misleading observed value functions.

The order for observation seasonality(p) and non-seasonality(q) is identified by autocorrelation function (ACF) and partial autocorrelation function (PACF) 

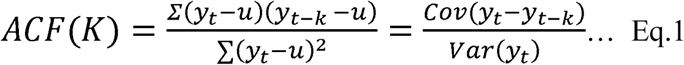

Where Yt is the original observed time-series value, Yt-k is a lagged of observed time-series, and ‘u’ is the mean value of observations, and k is the lag is the stationary observation characteristic. 

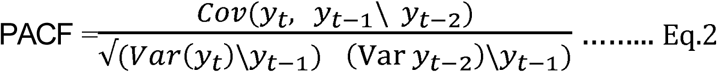

For our study, after the pre-processing method of data smoothening and testing the database for stationary and further for prediction modeling. Therefore, a multivariate database model for COVID-19 with different interaction methods was applied. We put the model with double differencing and as per lags for observed incidence (ARIMA(1,2,0)), mortality (ARIMA(0,2,2), and recover case (Brown’s method). ACF correlation is found more suited for the database, and therefore, the model we made for prediction of death, confirm, and recover variable separately is ARIMA (0,2,2) [14]. This ACF plot is understated with recovery cases and plotted in Figure 4. This will forecast observe value how the COVID-19 case causes prolonged influence, and in nearby date up to April 8, 2020, it will precisely show a graphical situation of upcoming days (Figure 4).

**Fig 1:**
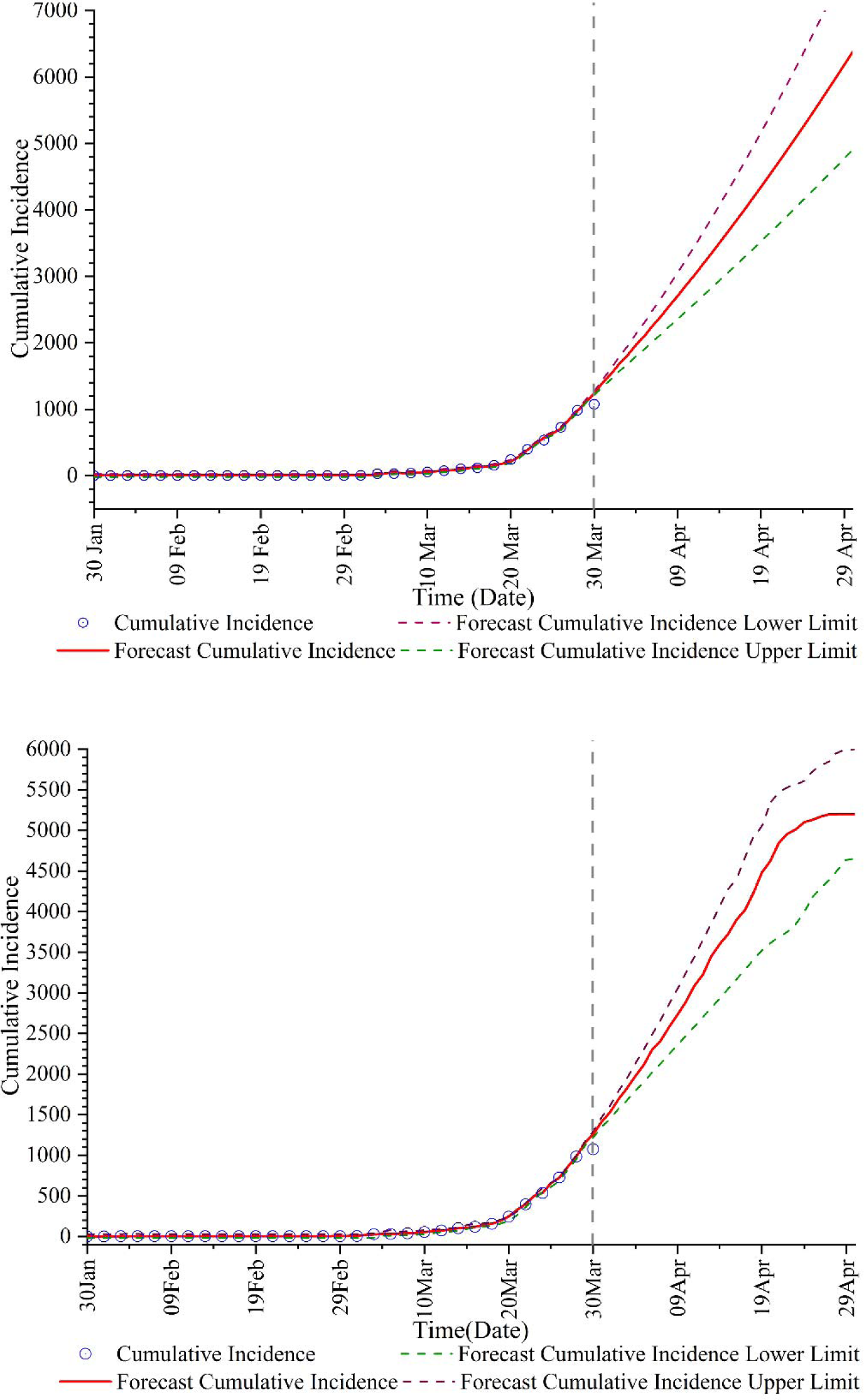
Thirty-day ahead ARIMA and Richard’s model forecasts of cumulative confirmed COVID-19 cases in India generated on 28 March, 2020.

**Fig. 2:**
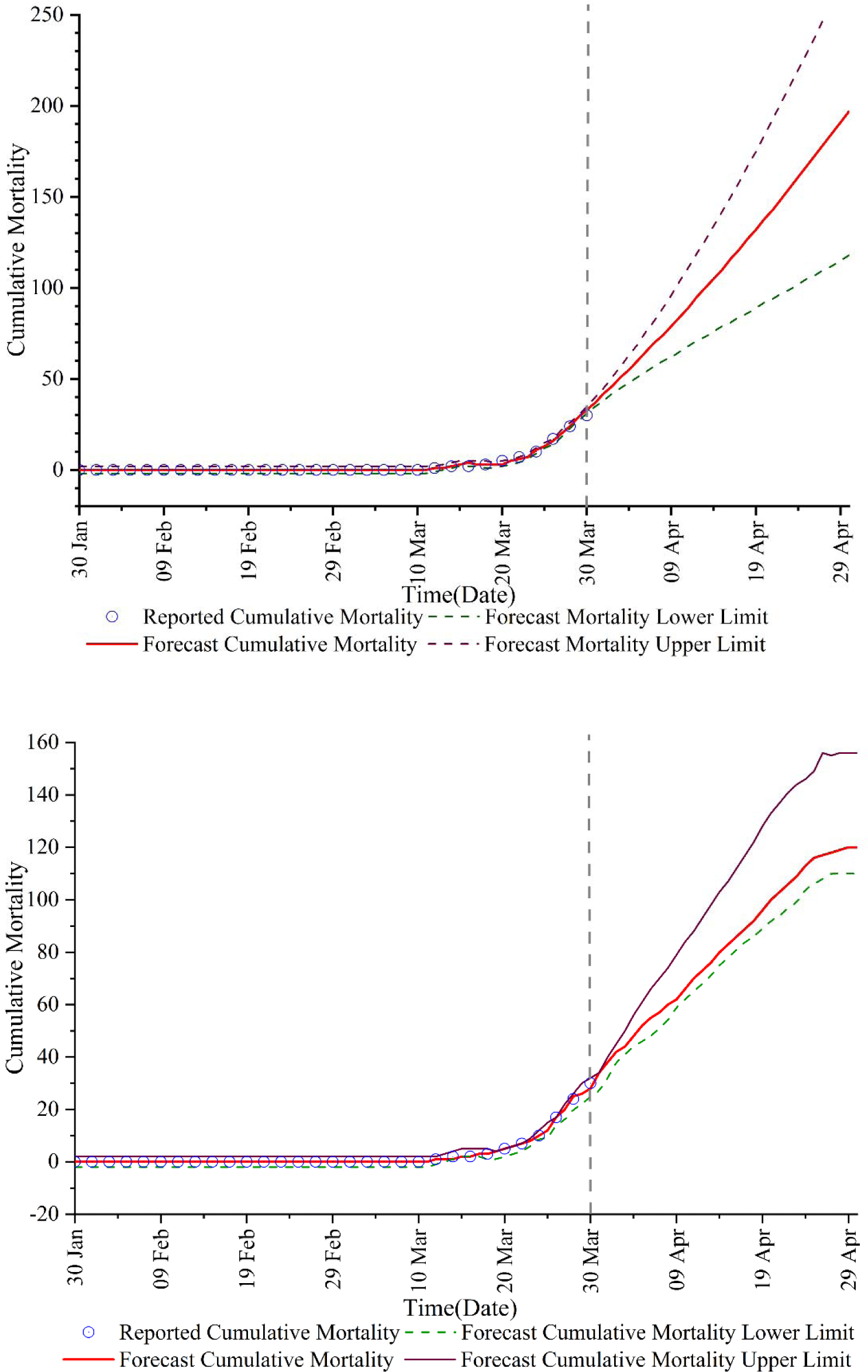
Thirty-day ahead ARIMA and Richerd’s model forecasts of cumulative death COVID-19 cases in India generated on 28 March, 2020.

**Fig. 3:**
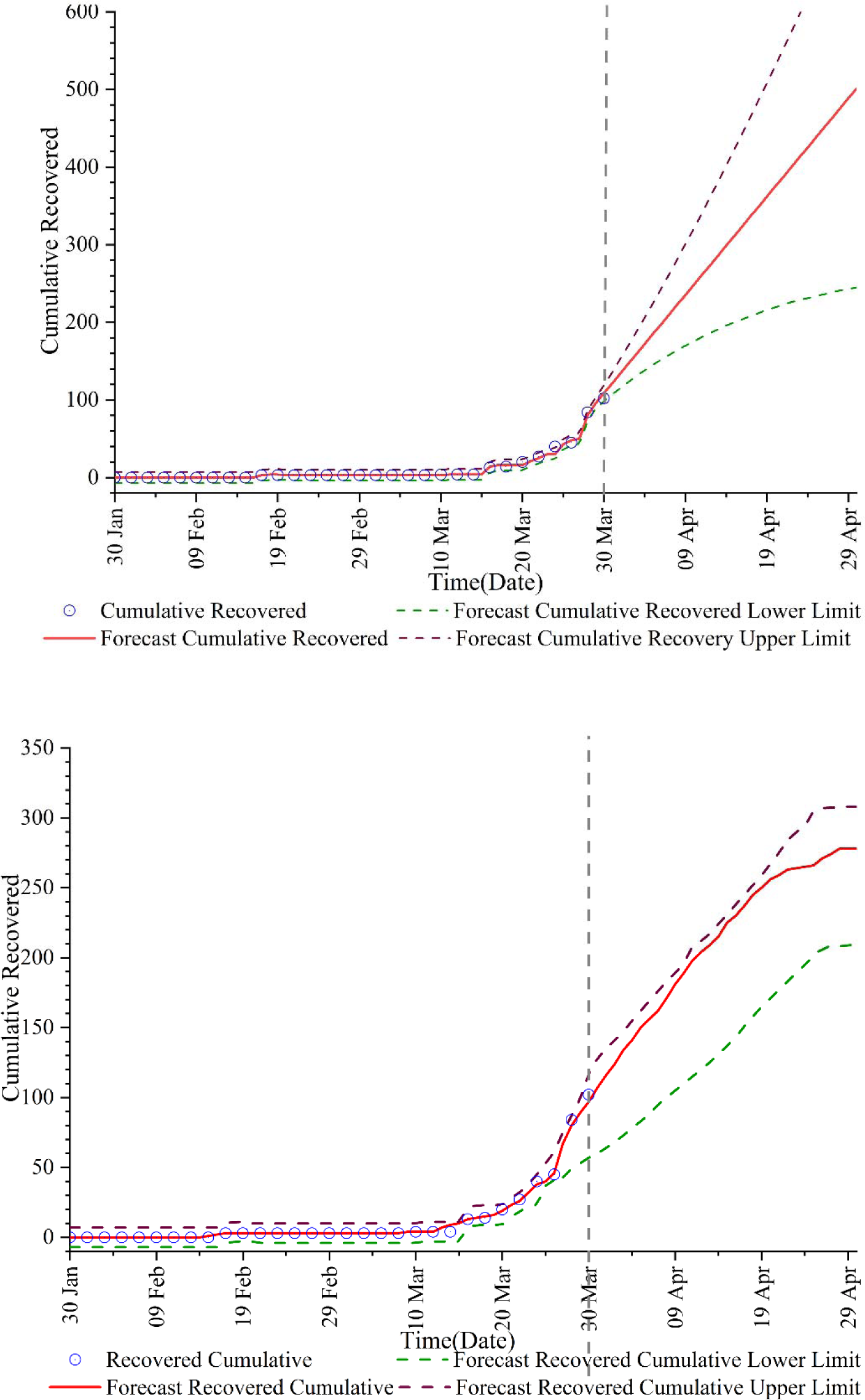
Thirty-day ahead ARIMA and Richerd’s model forecasts of cumulative recovered COVID-19 cases in India generated on 28 March 2020.

**Figure 4:**
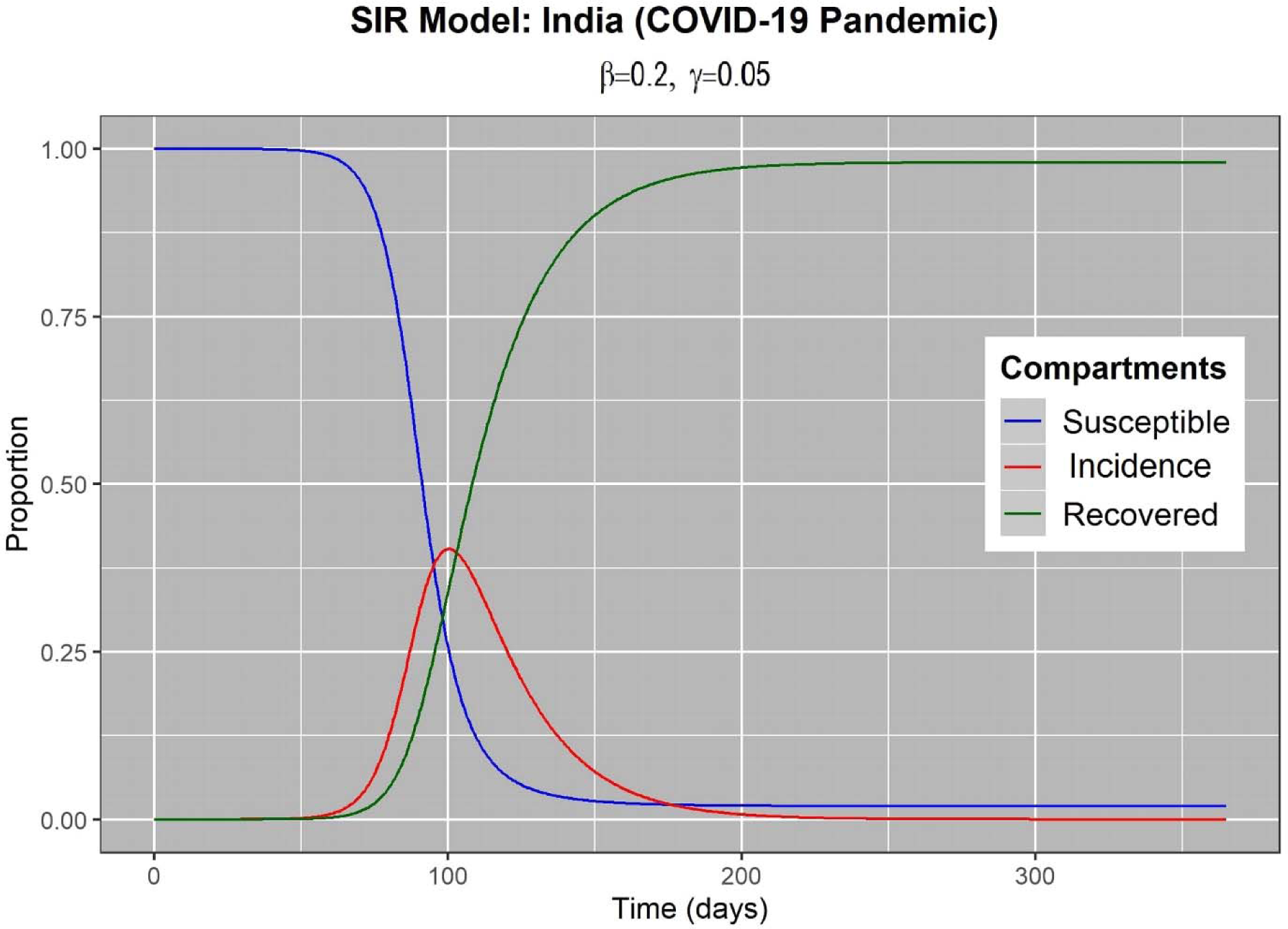
SIR representing the Incidence (Infected), (Susceptible) and Recovered cases in number of days’ time frame

### 3.5 Richerd’s Model

Richard’s is a non-linear sigmoidal function, a point of inflection occurring early in the adolescent stage, approaching a maximum value at an asymptote carrying capacity value (Fekedulegn et al., 1999). 

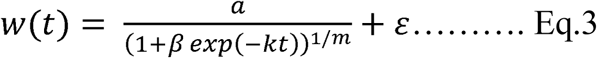

Where βgrowth range in the observed data, k is the observed data growth rate, t is the period, m is the slope of the observed data, and alpha(a) is the upper asymptote (upper value).

### SIR Model

The SIR model is one of the simplest and smartest compartmental models for epidemiology like COVID-19. The model is composed of three compartments that represent different categories of individuals within a population; the susceptible (S), infected (I), and removed (R). Hence it is called as SIR Model. Here susceptible is meant by people exposed to this COVID-19, infected comes after COVID-19 is confirmed in a person and removed means that either the person is recovered or death due to COVID-19. Mathematical it is determined concerning time, i.e., days here and given by: 

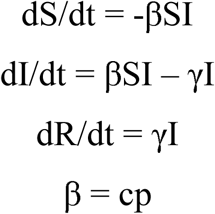

Where,

S – the proportion of susceptible individuals in total population

I – the proportion of infected individuals in total population

R – the proportion of removed individuals in total population

β – transmission parameter (rate of infection for susceptible-infected contact)

c – number of contacts each host has per unit time (contact rate)

p – the probability of transmission of infection per contact (transmissibility)

γ – recovery parameter (rate of infected transitioning to recovered)

This model is fundamental and has important assumptions. The first being the population is closed and fixed, in other words – no one is added into the susceptible group (no births), all individuals who transition from being infected to removed are permanently resistant to infection or are dead because of the disease, and there are no deaths. Second, the population is homogenous (all individuals are the same) and only differ by their disease state. Third, infection and that individual’s “infectiveness” or ability to infect susceptible individuals, coincides.

## MODEL Validation

We are presenting the short term forecast for the reported incidence, mortality, and recovered cases of nCOVID19 with the data incidence cases reported during the period of 30 January to 29 April 2020. The used two data model algorithm to predicted to know the outbreak of COVID19 during its next level of influence. We estimate the best-fit solution for each model using the nonlinear least-squares fitting, in which the test model provides the better goodness of fit.

The ARIMA and Richards both of the growth models validated based on Coefficient of determination(R^2^) desirable value should be higher, Root Mean Square Error (RMSE), its value should be lower, and Bayesian Information Criteria (BIC), the value should be lower. It provides the information for good model fit, and results are likely to be reliable. We observed that the ARIMA model was an excellent fit model.

|  | **Observation** | **$R^2$** | **RMSE** | **BIC** |
| --- | --- | --- | --- | --- |
| **ARIMA** | *Incidence* | 0.997 | 12.100 | 5.116 |
|  | *Mortality* | 0.973 | 0.833 | -0.174 |
|  | *Recovered* | 0.954 | 3.34 | 2.54 |
| **Richards** | *Incidence* | 0.91 | 1.340 | 6.116 |
|  | *Mortality* | 0.94 | 2.34 | 2.22 |
|  | *Recovered* | 0.944 | 2.34 | 5.54 |

### Prediction Modeling

Both models are non-linear ARIMA it predicts the straight line represents the COVID-19 outbreak will grow fastly, not represent any paucity, but its upper limit and lower limits are at the wider gap. In (b) Richards model, it is the nonlinear sigmoidal function, so it grows at a higher slope, then it tends to become a consistently higher level with any other statistical information; however, it upper and lower limits are also very close. The ARIMA model outperforms as a growth model in forecasts in the short term based on performance metrics that account for the certainty of the predictions the coverage at a 95% CI level.

The Cumulative incidence cases daily mapped, forecast short term next one month using more than two months of observations using ARIMA and Richards growth model (Figure.1). By the end of April 2020, the incidence of new cases is predicted to be 5200 (95% CI: 4650 to 6002) through ARIMA model (Figure1(a)), versus be 6378 (95% CI: 4904 to 7851) Richard’s model (Figure1(b)).

The mortality case estimated that there would be a total of 197 (95% CI: 118 to 277) deaths based on the ARIMA model (Figure2(a)) versus be 120 (95% CI: 110 to 156) for Richard’s model (Figure2(b)).

The drop-down in the recovery rates will reach around 501 (95% CI: 245 to 758) through ARIMA model(Figure3(a)), versus by the end of April 2020: 278 (95% CI: 116 to 380) for Richard’s model (Figure3(b))

SIR model represents the incidence (infected), suspectable and Recovered cases using ARIMA forecast data further spread of COVID tends cases to decrease in the epidemic incidence cases in India. After the above analysis and generation of models for prediction of COVID-19, it has been observed that the ARIMA model is more suited for prediction than comparing to Richard’s, and the output has come near to accuracy as validation. It can be identified as a very frightening future outcome; here, in this case, we predicted an overall analysis up to April 29, 2020, which defines fewer crises for India.

The ARIMA model shows a straight line with a very high slope in the cases on incidence and mortality and recovery case; however, Richard’s growth model in mid of forecast range it very great change in value and finally tends to become static for incidence and mortality and recovered cases. The ARIMA model the forecast limits are increased in the extensive limit as the time increases; however, Richard’s growth model limits in minimal range comparatively.

These preliminary results using ARIMA and Richard’s models can help guide future efforts to understand better the various spatial and social factors shaping sub-epidemic patterns for other infectious diseases.

## Conclusion

Through our investigation, we identified that by the end of April 2020, the incidence of new cases is predicted to be 5200 (95% CI: 4650 to 6002) through the ARIMA model versus be 6378 (95% CI: 4904 to 7851) Richard model. We estimated that there would be a total of 197 (95% CI: 118 to 277) deaths and drop down in the recovery rates will reach around 501 (95% CI: 245 to 758) by the end of April 2020. These estimates can help to strengthen the implementation of strategies to increase the health system capacity and enactment of social distancing measures all over India.

## Data Availability

All the data is obtained from the open-source

## Conflict of interest

None

## Notes

### Competing Interest Statement

The authors have declared no competing interest.

